# Immune memory in mild COVID-19 patients and unexposed donors from India reveals persistent T cell responses after SARS-CoV-2 infection

**DOI:** 10.1101/2020.11.16.20232967

**Authors:** Asgar Ansari, Rakesh Arya, Shilpa Sachan, Someshwar Nath Jha, Anurag Kalia, Anupam Lall, Alessandro Sette, Alba Grifoni, Daniela Weiskopf, Poonam Coshic, Ashok Sharma, Nimesh Gupta

**Author notes:** These authors have contributed equally to this work (Asgar Ansari, Rakesh Arya, Shilpa Sachan, Someshwar Nath Jha, Anurag Kalia). These authors have contributed equally to this work (Poonam Coshic, Ashok Sharma). Corresponding Author: Dr. Nimesh Gupta, Vaccine Immunology Laboratory, National Institute of Immunology, New Delhi -110067, India.

## Abstract

Understanding the causes of the diverse outcome of COVID-19 pandemic in different geographical locations is important for the worldwide vaccine implementation and pandemic control responses. We analyzed 42 unexposed healthy donors and 28 mild COVID-19 subjects up to 5 months from the recovery for SARS-CoV-2 specific immunological memory. Using HLA class II predicted peptide megapools, we identified SARS-CoV-2 cross-reactive CD4^+^ T cells in around 66% of the unexposed individuals. Moreover, we found detectable immune memory in mild COVID-19 patients several months after recovery in the crucial arms of protective adaptive immunity; CD4^+^ T cells and B cells, with a minimal contribution from CD8^+^ T cells. Interestingly, the persistent immune memory in COVID-19 patients is predominantly targeted towards the Spike glycoprotein of the SARS-CoV-2. This study provides the evidence of both high magnitude pre-existing and persistent immune memory in Indian population. By providing the knowledge on cellular immune responses to SARS-CoV-2, our work has implication for the development and implementation of vaccines against COVID-19.

## Introduction

The COVID-19 pandemic has evolved with variable trajectory in diverse geographical locations. Pre-existing immunity acquired from ‘common cold’ Human Coronaviruses (HCoVs) could have substantial implication in the immunological and epidemiological outcome of the pandemic. Because of the diverse geo-distribution and prevalence of HCoVs, there may be a varying impact of pre-existing immunity on the SARS-CoV-2 infection. Therefore, there is a considerable interest to understand the traits of pre-existing immunity and its impact on the virus spread and pathogenesis, disease outcome and the establishment of protective immunity in COVID-19.

In context of pre-existing immunity, the cross-reactive T cells are the focus of extensive investigations. Recent reports reveal the existence of cross-reactive CD4^+^ T cells in ∼20-50% of the individuals never been exposed to SARS-CoV-2 (Braun et al., 2020; Grifoni et al., 2020b; Le Bert et al., 2020; Mateus et al., 2020; Weiskopf et al., 2020). These cross-reactive CD4^+^ T cells are largely canonical memory cells and they may be the outcome of previous infections with many of the common cold HCoVs (Mateus et al., 2020). The cross-reactive memory CD4 T-cell subsets may lead to a favorable course of SARS-CoV-2 infection via direct anti-viral effects of CD4-CTL (Cytotoxic T Lymphocytes) or T helper cells, and also via establishing optimal germinal centers derived protective humoral immunity by follicular T helper cells. In fact, the cross-reactive immune memory to SARS-CoV-2 is limited to CD4^+^ T cells and more studies are required to understand the cross-reactivity from HCoVs in case of the humoral immunity (Premkumar et al., 2020; Wec et al., 2020; Yuan et al., 2020). Most of these studies are limited to the antibody analyses and there is no firm knowledge available for the cross-reactivity in the B cell pool. The Spike glycoprotein of SARS-CoV-2 is the major target of neutralizing antibodies (Premkumar et al., 2020; Walls et al., 2020). Particularly, antibodies targeting RBD display high neutralizing potential (Wajnberg et al., 2020) and shown to be predicative of survival (Secchi et al., 2020). However, there has been a concern over the decline of antibodies within first few months after SARS-CoV-2 infection (Long et al., 2020; Seow et al., 2020). Although, it’s not clear if this decline is gradual and if the similar decline exists in the memory pool of T cells and B cells.

In addition to SARS-CoV-2, the cross-reactive immunity acquired from the common cold HCoVs may have substantial impact on the immune response to COVID-19 vaccine. Therefore, there is an urgent need to understand the attributes of pre-existing immunity and quality of protective immune memory in COVID-19 across the diverse populations. In this study, we have examined the traits and stability of immune memory in unexposed donors and patients recovered from mild COVID-19. We show that the SARS-CoV-2 cross-reactive antibodies and CD4^+^ T cells exist in the unexposed donors, with Non-spike domains as the predominant target of CD4^+^ T cells in ∼66% of the individuals. Moreover, we also show that immunological memory to SARS-CoV-2 is detectable in mild COVID-19 patients up to 5 months (median ∼3 months) after recovery both in the CD4^+^ T cells and B cells. Interestingly, the durable immune memory in COVID-19 patients was highly targeted towards the Spike glycoprotein of the SARS-CoV-2. Our work provides the evidence of pre-existing reactivity and immune memory detectable in mild COVID-19 patients from the geographical location that is experiencing high burden of SARS-CoV-2 pandemic with an extremely low case fatality.

## Materials and Methods

### Ethics Statement

This study was approved by the Institutional review boards of the National Institute of Immunology and All India Institute of Medical Sciences, New Delhi, India. Informed consent was obtained from all subjects during the enrolment. For analyses in healthy individuals, buffy coat and plasma samples isolated from blood of healthy donors were collected from the blood bank in All India Institute of Medical Sciences, New Delhi, India.

### PBMC isolation

For all samples blood was collected in K3 EDTA tubes (COVID-19 donors) or EDTA coated blood bag (unexposed donors). Plasma was frozen at -80°C in multiple aliquots. PBMCs were isolated using Ficoll Paque Plus (GE Life Sciences) density gradient medium and cryopreserved in multiple aliquots in Fetal Bovine Serum (Gibco) containing 10% Dimethyl Sulfoxide (DMSO; Thermo-Fisher) and stored in liquid nitrogen until used in the assays. After revival, PBMCs were obtained with >80% viability, as accessed by acridine orange and propidium iodide double staining using the LUNA-FL (Logos Biosystems Inc., USA) automated cell counter. Details of the study population are provided in **Table 1**.

**Table 1.**
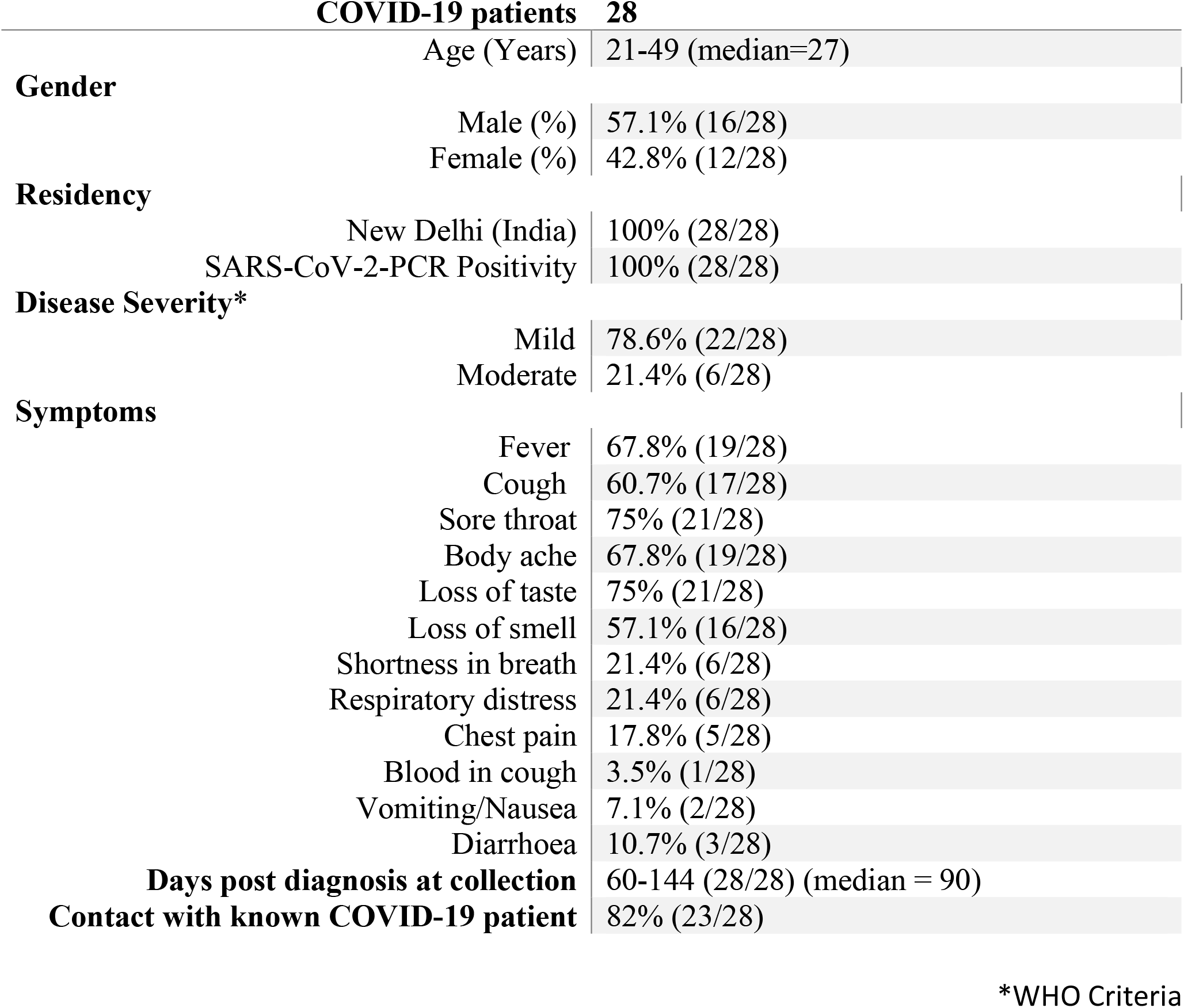
Characteristics of COVID-19 Patients.

### ELISA to detect SARS-CoV-2 specific IgG

ELISA plates (Nunc, Maxisorp) were coated with 100µl/well of SARS-CoV-2 full length Spike protein (Native Antigen, UK) and Nucleoprotein (Sino Biologicals) in PBS (pH 7.4) at the final concentration of 1µg/ml and incubated overnight at 4°C. After wash, the plates were blocked with blocking buffer (PBS containing 3% Skim milk and 0.05% Tween-20) and incubated at room temperature (RT) for 2 hours. Plasma samples were heat inactivated at 56^0^C for 1 hour. Plates were washed and 3-fold serially diluted heat inactivated plasma samples in dilution buffer (PBS containing 1% Skim milk and 0.05% Tween-20 in PBS) were added into the respective wells followed by incubation at RT for 1.5 hours. After incubation and wash, Goat anti-human IgG conjugated with Horseradish Peroxidase (HRP) (Southern Biotech) was added and plates were incubation at RT for 1 hour. The reaction was developed by adding o-phenylenediamine dihydrochloride (OPD) peroxidase substrate (Sigma-Aldrich) for 10 minutes in dark at RT. The reaction was stopped by adding 50µl/well of 2N HCl, followed by optical density (OD) measurement at 492 nm using MultiskanGO ELISA reader (Thermo-Fisher). The antigen coated wells that were added with sample diluent alone were used as blank to obtain the background OD values. For comparing the IgG titer in negative and COVID-19 recovered subjects, the Area Under Curve (AUC) was calculated for each specimen. The OD values obtained in test wells after subtracting the mean of background OD values were used for calculating the AUC, using a baseline of 0.05 for peak calculations. The positive response was defined as the value above the mean plus 3-times standard deviation of the lowest detected values, as in the case of reactivity with Spike protein, in all the tested samples from COVID-19 negative donors.

### ELISA to detect HCoV-OC43 and HCoV-NL63 specific IgG

The IgG reactivity to the Nucleoprotein of HCoV-OC43 and HCoV-NL63 was detected using in-house ELISA. The ELISA plates were coated with 100µl/well of HCoVs Nucleoprotein (Native Antigen, UK) in PBS at the final concentration of 1µg/ml and incubated overnight at 4°C. The plates were blocked and the reactivity was assessed in 1:100 diluted samples after incubation at RT for 1.5 hours. The Nucleoprotein-specific IgG were measured at 492 nm using the HRP-conjugated Goat anti-human IgG and OPD. The IgG reactivity was defined as the OD value in test wells after subtracting the mean of background OD values from blank wells.

### Virus neutralization assay

The neutralization potential of the antibodies in unexposed and COVID-19 recovered subjects was assessed by using the SARS-CoV-2 surrogate virus neutralization test (Tan et al., 2020). The test was performed following the manufacturer’s instructions (Genscript). Briefly, plasma samples were incubated with the RBD-HRP and the mixture was captured on the plate coated with human ACE2. The reaction was developed using the TMB substrate and the absorbance was measured at 450 nm using a microplate reader. The sample absorbance was inversely proportional to the titre of the anti-SARS-CoV-2 neutralizing antibodies. The percent neutralization was calculated using the formula: (1-OD value of sample/OD value of Negative Control) x 100%. The cut-off for the detection of SARS-CoV-2 neutralizing antibodies was determined by the manufacturer after validation with panel of confirmed COVID-19 patient sera and healthy control sera.

### Activation induced cell marker (AIM) assay for quantification of CD4^+^ T cells

Antigen-specific CD4 T-cell analysis was performed using the sensitive AIM assay (Havenar-Daughton et al., 2016; Reiss et al., 2017; Grifoni et al., 2020b). The PBMCs were stimulated for 24 hours in the presence of SARS-CoV-2 specific CD4 peptide megapools (MPs) (Spike protein: Spike; and remainder of the polyprotein: Non-spike) at 1 μ g/mL in 96-well U bottom plate in a total of 1×10^6^ PBMCs per well. For negative control, an equimolar amount of DMSO (vehicle) was added to unstimulated well. Stimulation with cytomegalovirus CD4 MP (CMV, 1 μ g/mL), or Staphylococcal Enterotoxin B (SEB) were included as positive controls. After 24 hours, cells were washed with 1 mL of PBS with 2% FBS (FACS buffer) and surface stained with antibody cocktail for 1 hour at 4° C in the dark; CD20, CD14, CD16, CD8a and fixable-viability dye coupled with APC eflour 780 in the dump channel, CD4-AlexaFluor 700 (RPA-T4), OX40-FITC (Ber-ACT35), CD137 PE Dazzle (4B4-1), CD45RA Brilliant Violet 785 (HI100) and CCR7 PE-Cy7 (3D12). Following the surface staining, cells were washed with FACS buffer and then fixed with freshly prepared 1% paraformaldehyde (Sigma Aldrich) for 30 minutes at 4°C in the dark. Cells were washed twice with FACS buffer and resuspended in FACS buffer before acquiring on a BD LSR Fortessa flow cytometer (BD Biosciences). Data were analysed using FlowJo 10.5.3. The positive response in the AIM assay was defined by setting up the limit of detection above the mean plus two-times of standard deviation of the response obtained in unstimulated conditions of all the unexposed and COVID-19 donors analysed. The frequency of responders to SARS-CoV-2 peptide pools was determined by applying the Fischer’s exact test on the AIM^+^ and AIM^-^ cells in unstimulated and peptide stimulated conditions. Stimulation Index (SI) was calculated by dividing the percentage of AIM^+^ cells after stimulation with peptide pools with the percentage of AIM^+^ cells derived from DMSO stimulation. The SI <1 was depicted as 1. The limit of positive stimulation index was defined as the median plus standard deviation of the lowest detected values, as in case of stimulation with Spike megapool, in COVID-19 unexposed donors.

### Activation induced cell marker (AIM) assay for quantification of CD8^+^ T cells

The antigen-specific analyses of CD8 T cells was performed using the AIM assay, similar to the above mentioned CD4^+^ AIM assay. The PBMCs were stimulated with the class I peptide megapool consist of 628 peptides from the whole virus proteome and split into two megapool, CD8-A and CD8-B containing 314 peptide each, as detailed previously (Grifoni et al., 2020a; Grifoni et al., 2020b). For negative control, an equimolar amount of DMSO (vehicle) was added to unstimulated well. Stimulation with Staphylococcal Enterotoxin B (SEB) were included as positive control. After stimulation, cells were washed and surface stained with antibody cocktail for 1 hour at 4°C in the dark; CD20, CD14, CD16, CD4 and fixable-viability dye coupled with APC eflour 780 in the dump channel, CD8-APC (RPA-T8), CD69-BV510 (FN50) and CD137 PE Dazzle (4B4-1). Following the surface staining the cells were washed, acquired and analysed as mentioned in the previous section of CD4-AIM assay. The positive response in the AIM assay was defined by setting up the limit of detection above the mean plus two-times of standard deviation of the response obtained in unstimulated controls of all the unexposed and COVID-19 donors analysed. Frequency of responders in unexposed and COVID-19 recovered subjects was determined using the combined data of CD8-A and CD8-B megapool after subtracting the background from the unstimulated controls.

### SARS-CoV-2 specific B cell ELISPOT

The antigen-specific memory B cells were measured in the cryopreserved PBMCs using polyclonal stimulation in RPMI 1640 in the presence of R848 (1 µg/mL) and IL-2 (10 U/mL) at cell density of 10^6^ PBMCs per well for 5 days. The Fluorospot plate (Mabtech) was charged with ethanol prior to the antigen coating. For antigen specific memory B cell analysis, plate was coated with SARS-CoV-2 full length Spike protein (Native Antigen, UK) and Nucleoprotein (Sino Biologicals) at concentration of 5µg/mL and incubated overnight at 4°C. As a control, for total memory B cell analysis, plates were coated with anti-human IgG, IgM, IgA coating antibodies (Mabtech) at concentration of 15µg/mL. Plates were washed and blocked with complete RPMI medium for at least 30 minutes at room temperature. Stimulated PBMCs were washed and seeded in complete RPMI at 0.5×10^6^ -1×10^6^ PBMCs per well for antigen-specific analysis and 20,000-50,000 cells for total B cell analysis. PBMCs were incubated at 37°C for 8 hours. Cells were discarded and plate was washed PBS. For detection of antibody secreting cell (ASC) spots, anti-human IgG-550, IgM-640 and IgA-490 detection antibodies (Mabtech) were added and plate was incubated for 2 hours at room temperature in dark. Plate was washed and fluorescence enhancer (Mabtech) was added to each well. ASC spots were detected on AID *vSpot* Spectrum Elispot/Fluorospot reader system using AID Elispot software version 7.x. As no spots were detected in wells without the antigen, presence of a spot >1 in the antigen-coated well was considered as a positive response. ASC counts were normalized to ASCs per million of PBMCs for all analyses.

### Statistical analysis

In all experiments, data are expressed as the mean±s.e.m. The significance of the differences between the groups was analysed with the two-sided Mann-Whitney test, Fischer’s exact test or Wilcoxon paired t-test as specified in the figure legends. P values < 0.05 were considered statistically significant. Statistical analyses were performed with the GraphPad Prism software version v8.

## Results

### Antibody response in unexposed donors and mild COVID-19 recovered patients with synchronous expansion of antibodies to HCoV-OC43

To investigate the quality and stability of immune memory in the COVID-19 patients we recruited 28 adult patients who had recovered from mild COVID-19 (Table 1). To explore the impact of cross-reactive immunity from ‘common cold’ coronaviruses we also utilized plasma samples and the peripheral blood mononuclear cells from 42 healthy blood donors collected prior to the pandemic during 2018-2019. The SARS-CoV-2 infection was diagnosed in all the recruited patients by viral PCR test. None of the patients required hospitalization and were quarantined with the mild-to-moderate manifestation of the disease. All the patients showed high titer IgG response to the full-length spike unlike unexposed donors that showed no evidence of spike-reactive IgG (Figure 1 A and C). SARS-CoV-2 nucleoprotein-reactive IgG was present in 15 of the 42 unexposed donors tested, and the titer was significantly higher in the mild COVID-19 recovered patients (Figure 1 B and D). We further analyzed neutralizing antibodies in 8 unexposed donors and 12 COVID-19 patients after ≥4 months of recovery. We observed the presence of highly effective neutralizing antibodies in all the patients after the long duration of recovery (Figure 1E). Because we observed cross-reactive antibodies to the SARS-CoV-2 nucleoprotein in unexposed donors, we examined the IgG reactivity to nucleoproteins from common-cold HCoV-OC43 and HCoV-NL63 as a representative betacoronavirus and alphacoronavirus, respectively. The IgG reactivity was present against both the HCoV-OC43 and HCoV-NL63 in almost all the unexposed donors. Interestingly, an increase in IgG reactivity in COVID-19 recovered patients was noted, but limited to the HCoV-OC43 (Figure 1F). Thus, the data suggest a detectable spike- and nucleoprotein-specific antibody response in the Indian patients recovered from mild disease, at least up to 5 months (median ∼3 months) post COVID-19 diagnosis. Surprisingly, COVID-19 patients showed an increase in IgG response against the HCoV-OC43 but not to the other common cold coronavirus tested, HCoV-NL-63, which may have less closely related Nucleoprotein to SARS-CoV-2 (Huang et al., 2020).

**Figure 1.**
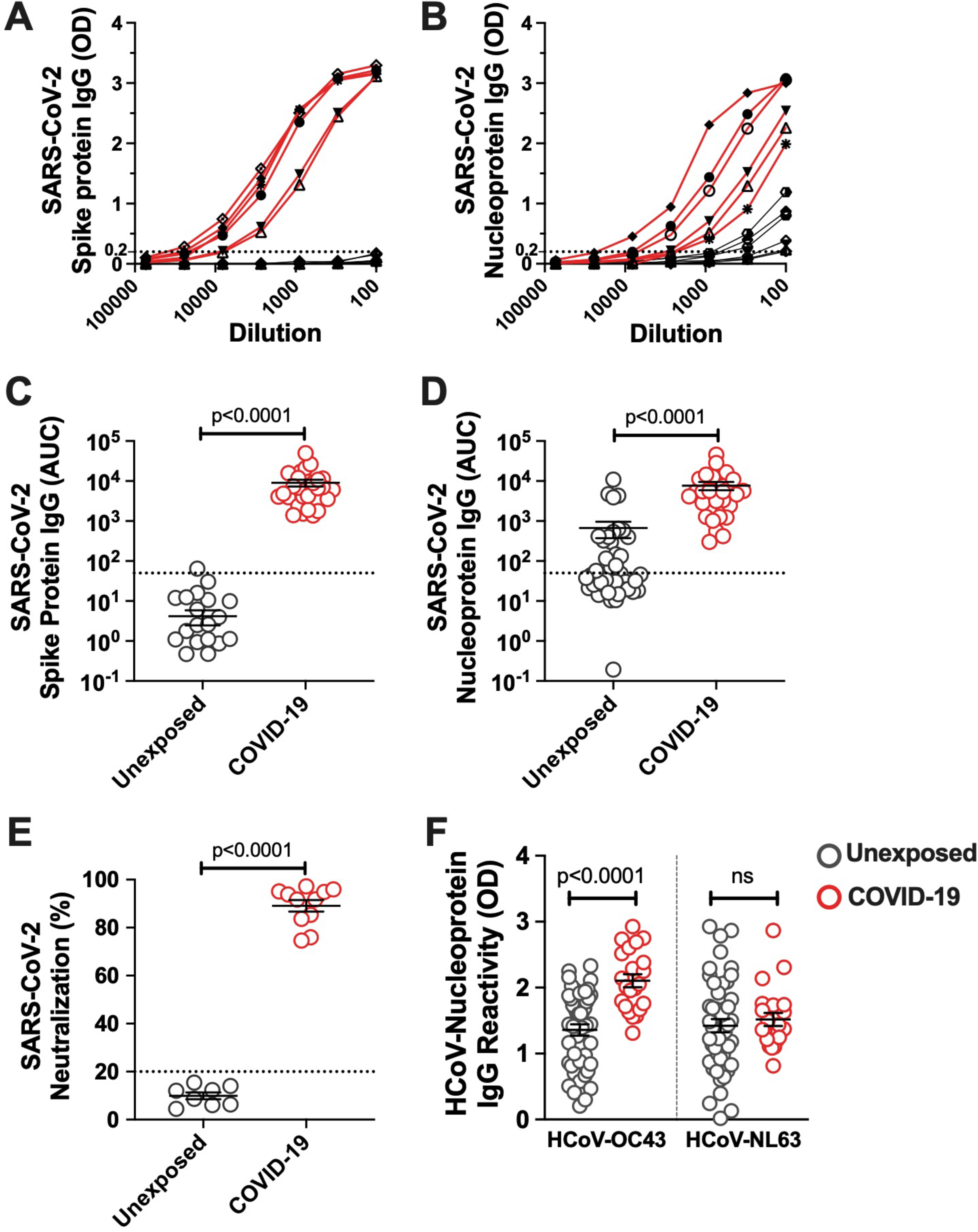
SARS-CoV-2 IgG response in pre-pandemic unexposed donors and individuals recovered from mild COVID-19. The IgG titre was measured in plasma sample from unexposed donors collected prior to pandemic and the COVID-19 patients up to 5 months of recovery by ELISA using the full length Spike protein and Nucleoprotein. The ELISA curves in serially diluted samples are shown from 6 representative unexposed donors (grey line) and COVID-19 cases (red line) for (**A**) Spike protein, (**B**) Nucleoprotein. Area under the curve (AUC) for ELISA quantitation of the IgG binding to (**C**) Spike protein, (**D**) Nucleoprotein for 42 unexposed donors and 28 COVID-19 cases inclusive of 6 representative donors of each group shown in panel A and B. (**E**) Neutralizing antibody quantitation in unexposed donors (n=8) and COVID-19 patients (n=12) after >4 months recovery measured using the SARS-CoV-2 surrogate virus neutralization test. (**F**) HCoVs Nucleoprotein antigen binding (expressed as OD) assessed by ELISA in unexposed donors (n=42) and COVID-19 recovered patients (n=28). Black bars indicate the geometric mean. Dotted line in panels A-E represent the cut-off of positivity. Statistical comparisons were performed by two-tail Mann-Whitney test. ns: non-significant.

### Robust SARS-CoV-2 specific CD4^+^ T-cell responses in unexposed donors and mild COVID-19 cases

CD4^+^ T cells are crucial for both the optimal quality of antibodies and anti-viral responses. Thus, we examined the CD4^+^ T cell reactivity in unexposed donors and the patients recovered from mild COVID-19. We measured the SARS-CoV-2 specific CD4^+^ T cells in the T cell receptor (TCR) dependent activation induced marker assay (Havenar-Daughton et al., 2016; Reiss et al., 2017). Here, we stimulated the PBMCs from 28 COVID-19 subjects and 32 unexposed healthy donors with the peptide megapool spanning the Spike domain (Spike) and the megapool covering the remainder of the SARS-CoV-2 genome (Non-spike) (Grifoni et al., 2020b; Mateus et al., 2020). A CMV megapool and the Staphylococcus Enterotoxin B (SEB) superantigen was used as the positive control, while DMSO was used as the negative control (Figure 2A and Supplementary Figure 1).

**Figure 2.**
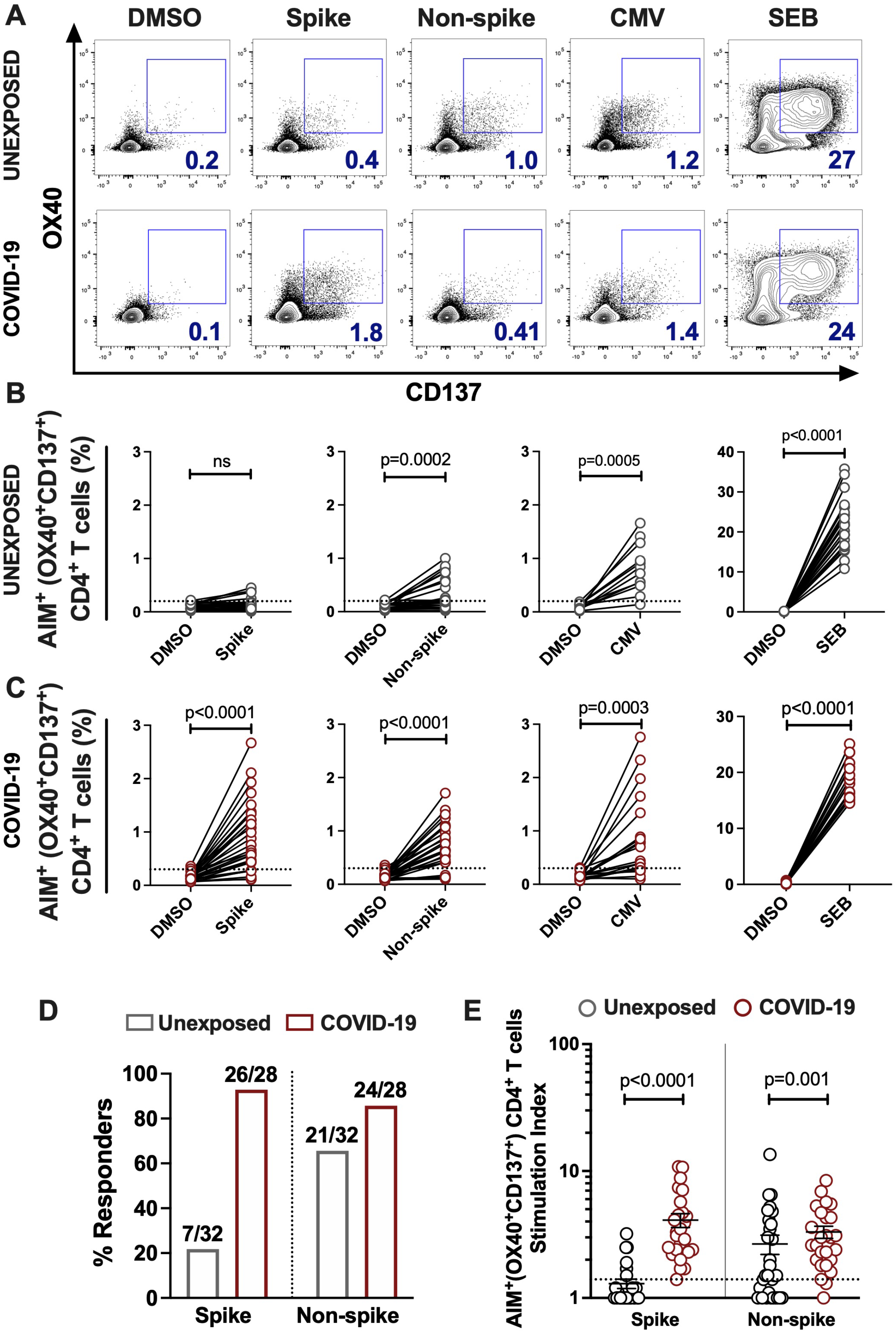
SARS-CoV-2-specific CD4^+^ T cells response in unexposed donors and recovered COVID-19 patients. The magnitude of SARS-CoV-2 specific CD4^+^ T cells was determined in PBMCs collected from unexposed donors (“Unexposed”, n=32) prior to pandemic and in COVID-19 patients (“COVID-19”, n=28) up to 5 months of recovery. The PBMCs were stimulated with the peptide megapool specific to Spike glycoprotein (Spike) or to the remainder of the SARS-CoV-2 polyprotein (Non-spike). DMSO was used as the negative control, and CMV peptide megapool and SEB were used for positive stimulation controls. **(A)** Representative FACS contour plots of unexposed and COVID-19 patient in stimulation conditions of DMSO, Spike peptide megapool, Non-spike peptide megapool, CMV and SEB. Paired graphs depicting the reactivity of AIM^+^ (OX40^+^CD137^+^) CD4^+^ T cells between the negative control (DMSO) and antigen-specific stimulation in (**B**) Unexposed donors (**C**) COVID-19 patients. **(D)** Frequency of responders to Spike and Non-spike peptide pools in unexposed and COVID-19 recovered subjects as determined by the Fischer’s exact test. The value on bars denote the number of responders/total number of donors tested. **(E)** Stimulation index quantitation of the AIM^+^ (OX40^+^CD137^+^) CD4^+^ T cells in Unexposed versus COVID-19 cases analysed in the same samples as in panel B and C. Black bars indicate the geometric mean. Dotted line in panels B, C and E represent the limit of detection. Statistical comparisons were performed by (B-C) Wilcoxon paired t-test and (E) two-tail Mann-Whitney test. ns: non-significant.

A total of 7 out of 32 unexposed donors were associated with marginal frequency of SARS-CoV-2 spike-reactive AIM^+^ (OX40^+^CD137^+^) CD4^+^ T cells with an insignificant increase over the DMSO control (Figure 2B and 2D). Interestingly, 21 out of 32 unexposed donors robustly responded to the peptide megapool covering the Non-spike domains of virus with a significantly higher frequency of AIM^+^CD4^+^ T cells over the DMSO control (Figure 2B; DMSO vs Non-spike pool, P=0.0002 and Figure 2D). The unexposed donors consistently responded to the CMV peptide megapool and the SEB superantigen significantly over the DMSO control (Figure 2B; DMSO vs CMV pool, P=0.0005; DMSO vs SEB, P<0.0001). The COVID-19 recovered patients showed robust activation and detectable SARS-CoV-2-specific CD4^+^ T cells in response to the Spike (26/28; 93%) (Figure 2C; DMSO vs Spike megapool, P<0.0001 and Figure 2D) and to the Non-spike peptide pool (24/28; 86%) (Figure 2C; DMSO vs Non-spike megapool, P<0.0001and Figure 2D). Like unexposed donors, COVID-19 patients readily responded to the CMV peptide pool and SEB stimulation (Figure 2C; DMSO vs CMV pool, P=0.0003; DMSO vs SEB, P<0.0001). Moreover, no significant correlation was observed between the frequency of Spike-specific CD4^+^ T cells and the days from symptoms onset in convalescent patients (Supplementary Figure 2A). Next, we measured the stimulation index of antigen specific stimulations over the unstimulated DMSO control to quantify CD4^+^ T cell reactivity in case of pre-existing immunity and in long-term post recovery from COVID-19. We observed a remarkably higher frequency of Spike-specific memory CD4^+^ T cells in recovered patients than the unexposed donors (Figure 2E; Unexposed vs COVID-19, P<0.0001). Surprisingly, higher magnitude of Non-spike reactive CD4^+^ T cells were also present in the unexposed donors as in recovered COVID-19 patients (Figure 2E; Unexposed vs COVID-19, P=0.001). Next, we determined the memory phenotype of the CD4^+^ T cells responding to the spike and non-spike peptide megapools (Supplementary Figure 3A). Both the central memory and effector memory compartments were mainly populated in antigen-specific CD4^+^ T cells, with no significant difference in the proportion specific to spike or non-spike genome of SARS-CoV-2 (Supplementary Figure 3B).

We further utilized the Class I peptide megapool to measure the SARS-CoV-2 specific CD8^+^ T cells in unexposed and recovered COVID-19 patients (Supplementary Figure 4A). The megapool consist of 628 peptides spanning the whole virus proteome and split into two pools, CD8-A and CD8-B, containing 314 peptides each (Grifoni et al., 2020b). The minimal CD8^+^ T cell responses were detected only in the stimulation with CD8-A megapool, which consist of spike epitopes including the epitopes of other proteins (Supplementary Figure 4 B-C). The unexposed donors and COVID-19 patients consistently responded to the SEB superantigen significantly over the DMSO control (P<0.0001; Supplementary Figure 4 B-C). By combining the responses in both the megapool CD8-A and CD8-B, total CD8^+^ T cell responses were detected in 2 of 18 unexposed donors and 4 of 18 recovered COVID-19 patients (Supplementary Figure 4 D-E).

Altogether, the antigen-specific T cell analyses suggest predominant and widespread CD4^+^ T cells responses over the CD8^+^ T cells in both the unexposed and recovered mild COVID-19 patients. There was a minimal presence of Spike-specific CD4^+^ T cells in unexposed donors with a remarkably high magnitude in case of recovery from mild COVID-19. Interestingly, almost similar magnitude of non-spike specific CD4^+^ T cells are present in majority of the unexposed and COVID-19 recovery patients. Detection of Spike-specific memory CD4^+^ T cells several months after infection is encouraging for the efforts focusing on SARS-CoV-2 Spike protein as a vaccine candidate.

### High magnitude Spike-specific B cells in mild COVID-19 recovered subjects

Because the mild COVID-19 patients showed robust Spike-specific CD4^+^ T cells reactivity, we examined if a similar finding would extend to SARS-CoV-2 B cell responses. Thus, utilizing SARS-CoV-2 Spike protein and Nucleoprotein (representative of Non-spike domains), we analyzed the frequency of each isotype-specific antibody secreting B cell population in unexposed subjects and the COVID-19 patients up to 5 months of recovery from mild disease (Figure 3A). The magnitude of IgG antibody secreting cells (ASC) was the highest among three subsets analyzed, as seen in the patients ∼4 weeks after recovery (Juno et al., 2020). Surprisingly, all the patients showed significant 6-fold higher Spike-specific IgG-ASC over the ASCs specific to Nucleoprotein (Figure 3B; Count/10^6^ PBMCs: Spike -780±84, Nucleoprotein - 131±35; P<0.0001). The IgG-ASCs were also detected in around 6 (Spike-specific) and 14 (Nucleoprotein-specific) of the 28 unexposed subjects, with the substantially lower frequency than the COVID-19 patients (Figure 3B). Although the frequency of Nucleoprotein- and Spike-specific IgM-ASCs were significantly higher than the unexposed subjects, it was not significantly different in the COVID-19 recovered patients (Count/10^6^ PBMCs: Spike - 427±70, Nucleoprotein - 463±76) (Figure 3C). Plasma cells secreting IgA were present in the least frequency in COVID-19 recovered patients and was only detected in the 8 (Spike-specific) and 10 (Nucleoprotein-specific) of the 28 unexposed subjects. Unlike Spike-specific IgA-ASCs that were detected in all the recovered patients, the Nucleoprotein specific IgA-ASCs were present in 13 of the 18 donors tested. However, like IgG-ASCs, Spike-specific memory IgA-ASCs were present in 2-fold higher frequency than the Nucleoprotein-specific cells in COVID-19 patients (Figure 3D; Count/10^6^ PBMCs: Spike - 65±12, Nucleoprotein - 33±9; P=0.009).

**Figure 3.**
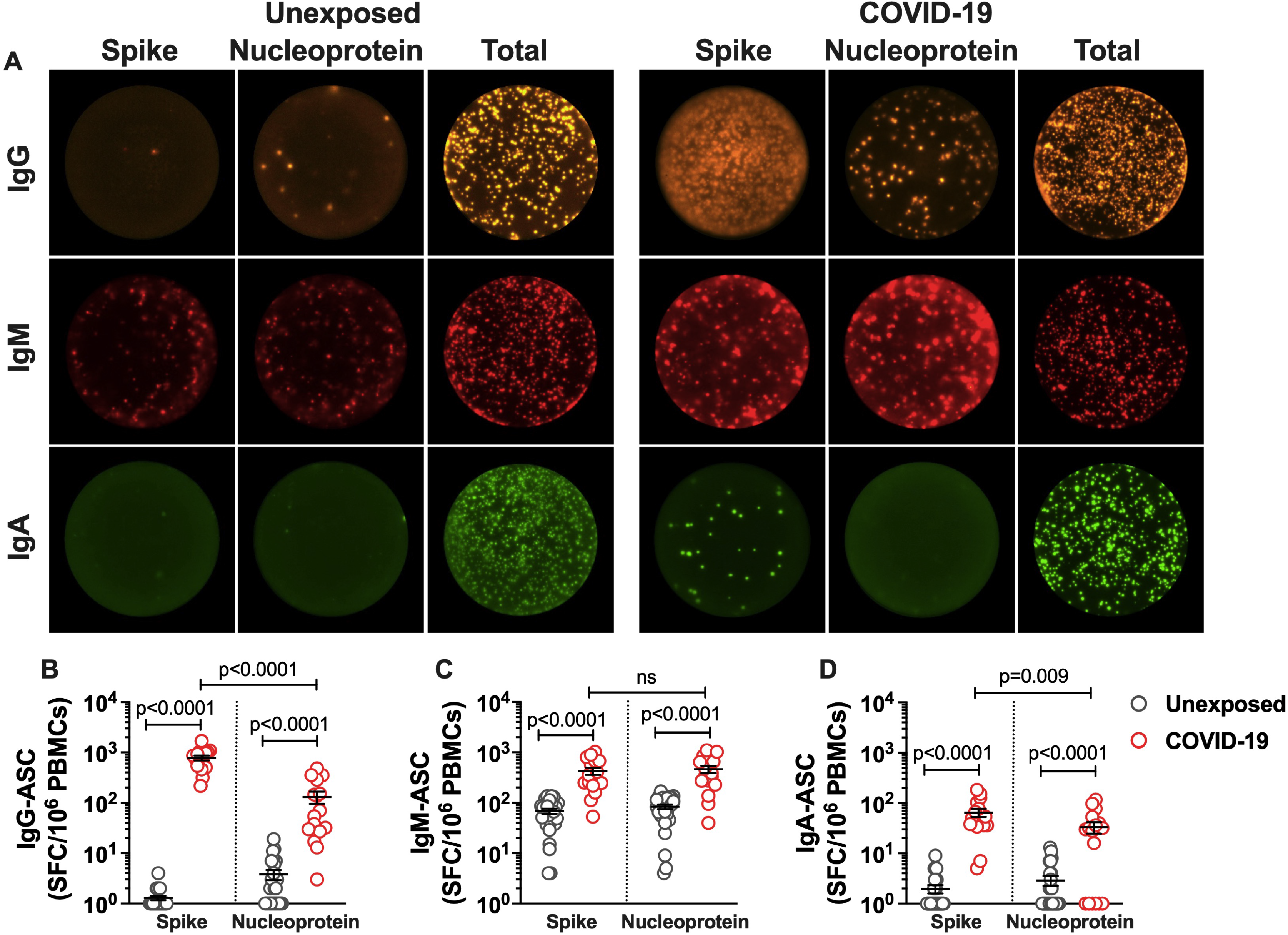
SARS-CoV-2-specific memory B cells in recovered COVID-19 patients. The frequency and isotype distribution of antibody secreting B cells (ASC) was measured in the unexposed subjects (“Unexposed”, n=28) prior to pandemic and in patients (“COVID-19”, n=18) up to 5 months of recovery from mild COVID-19. The memory B cells in PBMCs were polyclonally stimulated before measuring the frequency of SARS-CoV-2 Spike glycoprotein- and Nucleoprotein-specific IgG, IgM and IgA antibody secreting cells in Fluorospot assay. **(A)** Representative images of IgG, IgM and IgA secreting B cells in Unexposed subject and recovered COVID-19 patient. Graphs depicting the magnitude of antibody secreting B cells specific to the SARS-CoV-2 Spike glycoprotein and Nucleoprotein (expressed as spot forming cells (SFC) in 10^6^ PBMCs) for (**B**) IgG-ASC (**C**) IgM-ASC and (**D**) IgA-ASC, in Unexposed subjects (grey circle) and COVID-19 patients (red circle). For log scale, the spot count of less than one is depicted as 1. Black bars indicate the geometric mean. Statistical comparisons were performed by two-tail Mann-Whitney test. ns: non-significant.

Altogether, these results indicate the existence of high magnitude IgG secreting cells in the antigen specific B-cell pool of mild COVID-19 patients. A small fraction of unexposed subjects showed cross-reactive ASCs present in a very low frequency. Like in the case of CD4^+^ T cells, a significant number of B cells is found in long-term after recovery from mild COVID-19, targeted towards the Spike protein of SARS-CoV-2.

## Discussion

Here, we report the extent of pre-existing immunity and immune memory in individuals from 2 to 5 months (median ∼3 months) after the diagnosis of COVID-19. The existence of high titer Spike- and Nucleoprotein-specific IgG after several months post-infection indicates persistent antibody response in mild disease. Our observation is consistent with the recent reports where no decline was observed in antibodies to SARS-CoV-2 within 4 to 5 months of the COVID-19 diagnosis (Gudbjartsson et al., 2020; Wajnberg et al., 2020). This is important for the vaccine development as the mild disease may provide the crucial knowledge for generating a long-term sustainable antibody response.

In our cohort, SARS-CoV-2 Spike cross-reactive antibodies were not detected in the unexposed donors’ samples. This may be due to highly divergent Spike of SARS-CoV-2 than the seasonal coronaviruses (Forni et al., 2017). By contrast, almost 35% of the unexposed adult donors showed the existence of SARS-CoV-2 Nucleoprotein reactive antibodies. Unlike Spike protein, Nucleoprotein antibodies are more cross-reactive within the subgroups of HCoVs (Agnihothram et al., 2014) and it’s likely that the adult population in India has been exposed to common cold HCoVs as frequently as in the case with children and adolescents (Ng et al., 2020). The apparent nucleoprotein cross-reactivity seems to best correlate with the HCoV-OC43 Nucleoprotein-specific antibodies as increased titers associated with SARS-CoV-2 infection were observed with HCoV-OC43, a representative betacoronavirus used in this study. This may be due to more conserve Nucleoprotein immunodominant regions within the same family of betacoronaviruses (Meyer et al., 2014). Although, it’s not clear if the similar or the unique epitopes of HCoV-OC43 are associated with this observed expansion. Indeed, the high titer Nucleoprotein targeting antibodies in unexposed donors and in long-term after recovery warrants detailed study to identify their implication in the SARS-CoV-2 pathogenesis and the disease outcome.

There is no information available on the pre-existing cross-reactive T cells in the Indian population. We show that the cross-reactivity to SARS-CoV-2 as well as the memory responses are mostly associated with the CD4^+^ T cells with a minor contribution from CD8^+^ T cells. The minimal contribution of CD8^+^ T cells among the cross-reactive T cells was also observed in other cohorts (Grifoni et al., 2020b). The lack of SARS-CoV-2 specific CD8^+^ T cells in majority of the patients recovered from mild disease may be due to the poor stability or due to an inefficient establishment of the memory CD8^+^ T cells. Future investigations in different disease outcome across diverse populations are necessary to understand the implication of CD8^+^ T cells in SARS-CoV-2 pathogenesis. Our observation of poorly detected SARS-CoV-2 Spike-reactive CD4^+^ T cells in ∼20% of unexposed donors is consistent with the findings in the USA and the German cohorts (Braun et al., 2020; Grifoni et al., 2020b). However, higher frequency of SARS-CoV-2 Non-spike specific CD4^+^ T cells were observed in ∼66% of donors prior to the pandemic as compared to ∼50% in USA and the Singapore cohort (Grifoni et al., 2020b; Le Bert et al., 2020). In the Non-spike peptide megapool, the Nucleoprotein is the mainly targeted structural domain by the cross-reactive CD4^+^ T cells (Mateus et al., 2020). Because of substantial homology of Nucleoprotein between common cold HCoVs and SARS-CoV-2 and due to high prevalence of related common cold HCoVs, as supported by the IgG reactivity, it’s plausible that a higher extent of Nucleoprotein cross-reactive CD4^+^ T cells are present in our cohort of unexposed donors. Certainly, in-depth analyses in the unexposed donors are necessary to reveal if the prevalence and frequency of common cold HCoVs defines the targets of cross-reactivity to SARS-CoV-2 genome.

The cross-reactive CD4^+^ T cells might not be implicated solely in terminating the virus infection however they may limit the virus burden and reduce the course of symptomatic infection leading to lower incidences of severe disease (Lipsitch et al., 2020). This is particularly interesting in context of the high frequency Nucleoprotein-specific cross-reactive CD4^+^ T cells. Nucleoprotein is the first and most abundantly produced multifunctional protein in the virus infected cells (de Wit et al., 2016). The pre-existing cross-reactive CD4^+^ T cells may limit the virus spread by cytolysis of the infected cells that are displaying the processed Nucleoprotein on their surface early in the infection. By controlling the virus spread and reducing the virus burden, pre-existing cross-reactive CD4^+^ T cells might be implicated in providing a competitive window to the host to initiate an optimal protective immune response against the SARS-CoV-2.

Interestingly, high magnitude of Spike- and remainder of the genome CD4^+^ T cell responses are present in the patients long after recovery from mild COVID-19. However, unlike Spike-specific CD4^+^ T cells that show a substantially higher magnitude over the cross-reactive T cells in unexposed donors, the non-spike specific memory CD4^+^ T cells are associated with a lesser increase after COVID-19. Future studies may reveal if there is an influence of cross-reactive memory T cells on de novo generation of non-cross-reactive clones targeting the Non-spike domains (Brehm et al., 2002; Johnson et al., 2016) or this diverse outcome is due to an immunodominance of CD4^+^ T cells targeting the highly immunodominant Spike glycoprotein leading to de novo expansion of Spike-specific CD4^+^ T cells and outcompeting the expansion of CD4^+^ T cells targeting low frequency T-cell epitopes present in Nucleoprotein and the remainder of the genome (Olson et al., 2016; Grifoni et al., 2020a; Mateus et al., 2020). Interestingly, in the similar lines, higher magnitude of Spike-specific IgG and IgA secreting B cells over Nucleoprotein-specific B cells further supports the notion of targeted and persistent

immune response to a highly immunodominant Spike glycoprotein of SARS-CoV-2 in mild disease. The IgM secreting B cells were present at lower frequency than IgG secreting B cells in COVID-19 recovery. However, these IgM secreting B cells were also detected in all the unexposed subjects. It’s plausible that these IgM secreting B cells were not antigen-selected and developed in absence of a productive germinal center reaction during previous infection with the closely related human common cold coronaviruses (Bohannon et al., 2016). More studies in longitudinal prospective cohort are necessary to reveal the implication of pre-existing IgM secreting B cells in the SARS-CoV-2 pathogenesis and to determine if lower frequency of IgM secreting B cells is due to a poor stability in long-term or it’s due to limited de novo generation in response to the SARS-CoV-2.

The limitations in our study include the sample size and the longitudinal sampling to probe the stability in immunological memory. As this study was of exploratory nature, the 28 long-term recovery samples represent the recruitment in a reasonable timeframe. In fact, the sample size was sufficient to determine the existence of cross-reactive CD4^+^ T cells and to reveal the persistence of memory CD4^+^ T cells in several months after recovery from COVID-19. Besides, the predicted epitopes utilized in this study to examine the CD4^+^ T cells may not cover the responses to all the epitopes in viral genome. However, these predicted peptide pools cover most of the immunodominant epitopes and provide an opportunity to detect the virus-specific CD4^+^ T cells in limitedly available patient blood sample. Certainly, further studies in long-term after recovery in a larger longitudinal cohort will be helpful in defining the breadth and durability of SARS-CoV-2 reactive memory CD4^+^ T cells. Moreover, it will be very important to determine if similar characteristics of memory CD4^+^ T cells exist in the recovery from different outcomes of disease from the asymptomatic to the severe COVID-19. While our work was in review, the knowledge on multiple virus variants emerged in the literature. Because the peptide pools used in our study originates from the reference strain of SARS-CoV-2 (GenBank: MN908947), the current analyses do not provide the information on reactivity of T cells to the mutated epitopes in recent virus variants. However, this remains an area of the future study to determine the capability of memory T cells established against the previously circulating virus in responding to the respective mutated epitopes in the recently emerged virus variants.

In summary, we show that the individuals recovered from mild disease display a response detectable several months after recovery in two crucial arms of protective immunity - CD4^+^ T cells and B cells. We also show the existence of pre-existing immunity in the unexposed donors, which is predominantly associated with the non-spike part of the genome of SARS-CoV-2. Although the cross-reactive T cells are present against both the spike and non-spike epitopes, the magnitude of cross-reactive CD4^+^ T cells targeting the non-spike epitopes is extremely high in our cohort. Indian continent has seen high burden of the COVID-19 incidences; however, the case fatality rates are extremely low. Whether high magnitude of cross-reactive CD4^+^ T cells are contributing to this less severe outcome needs to be addressed in the prospective cohort before and after COVID-19. The knowledge on implication of cross-reactive CD4^+^ T cells in the disease outcome and in establishment of immunological memory is crucial for the development and implementation of COVID-19 vaccines.

## Data Availability

Not Applicable

## Acknowledgements

We are thankful to all the patients for generous support in this study, and Mr. Neeraj, Mr. Jagat Singh (AIIMS) and Mr. Sudipta Das (NII) for technical support. This work was supported by Science and Engineering Research Board, DST grant IPA/2020/000077 (to NG, AS, PC), Biotechnology Industry Research Assistance Council, DBT grant BT/COVID0010/01/20 (to NG). Further support provided from NIH contract 75N9301900065 (to A.S, D.W) and NIH grant U01 (U01AI141995-03) to A.S.

## Author contributions

R.A, J.L, A.S, and P.C: Enrolled and categorized subjects, collected samples and provided clinical information. A.A., S.S., S.N.J, A.K: performed experiments. A.S, A.G, and D.W: Contributed essential material. N.G.: Conceived and supervised the study, analysed the data and wrote the manuscript. A.S and D.W: critically reviewed the manuscript.

