## Supplementary Figures 1 to 4 for "Immune memory in mild COVID-19 patients and unexposed donors from India reveals persistent T cell responses after SARS-CoV-2 infection"

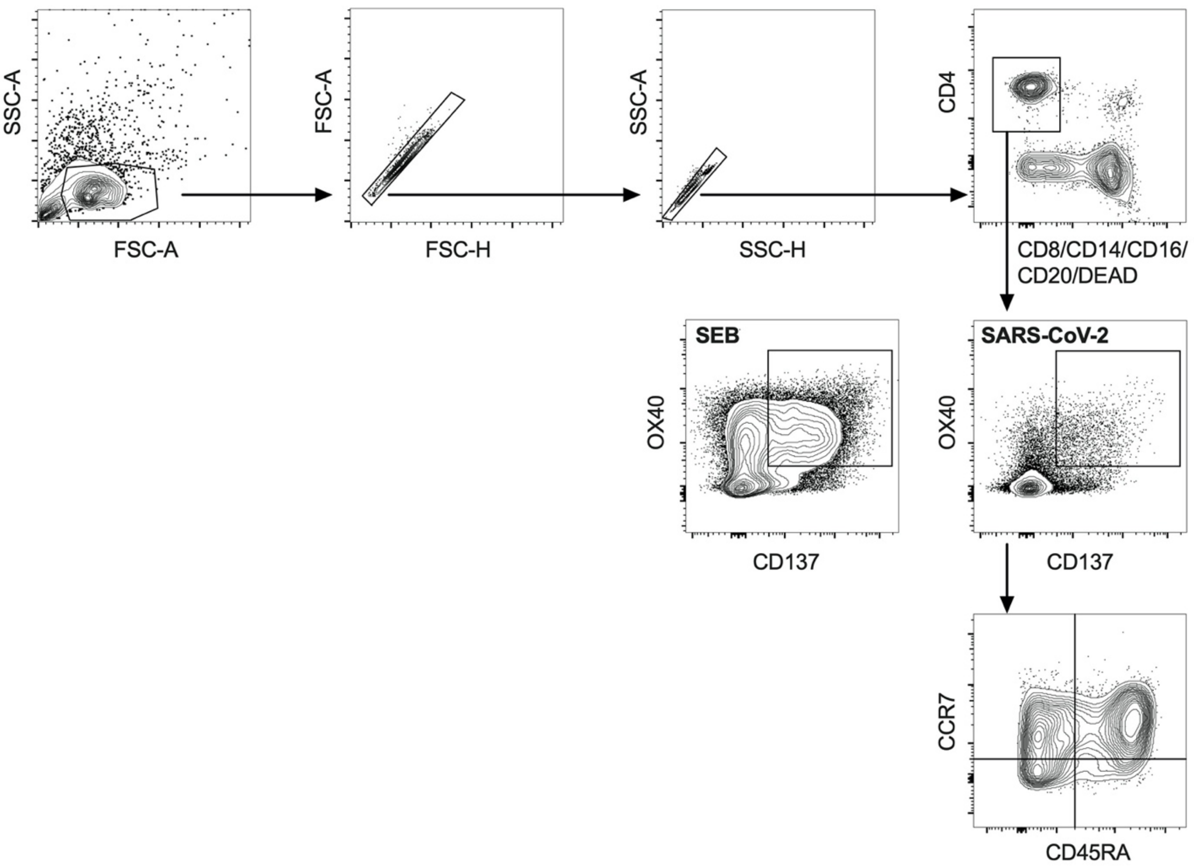


**Supplementary Figure 1. Flow cytometry Gating Strategy.** The example gating strategy for analysing the OX40$^{+}$CD137$^{+}$ (AIM$^{+}$) CD4$^{+}$ T cells in the PBMCs from unexposed donors and the recovered COVID-19 patients.

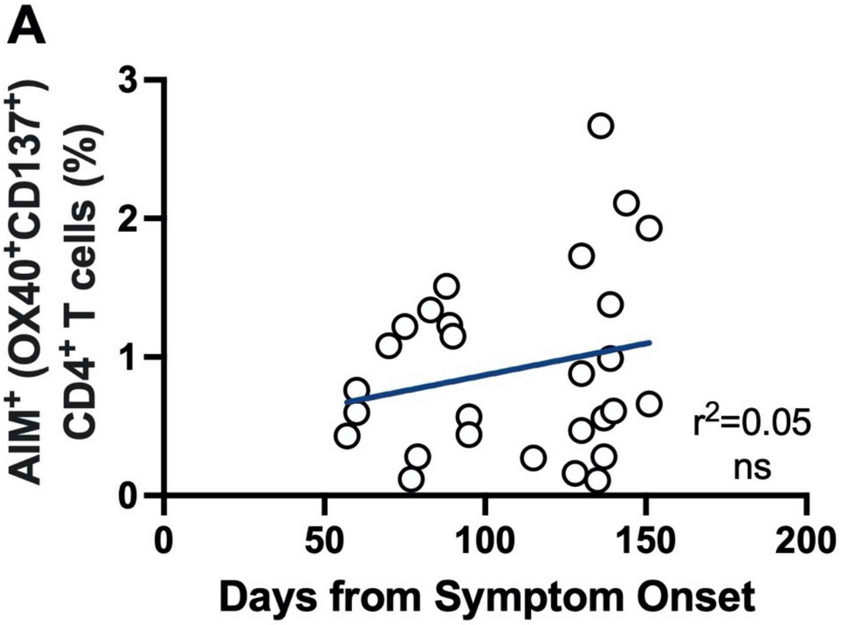


**Supplementary Figure 2. Correlation of SARS-CoV-2 Spike-specific CD4$^+$ T cell response with the time from symptoms onset.** The frequency of SARS-CoV-2 specific CD4$^+$ T cells was determined in the subjects recovered from COVID-19. The PBMCs were stimulated with the peptide megapool specific to Spike glycoprotein (Spike) and antigen-specific cells were determined using the AIM assay. (**A**) Correlation of the frequency of AIM$^+$ Spike-specific CD4$^+$ T cells with the days from symptoms onset in recovered patients. Each circle represents a COVID-19 subject. Statistical analysis was performed by simple linear regression. ns: non-significant.

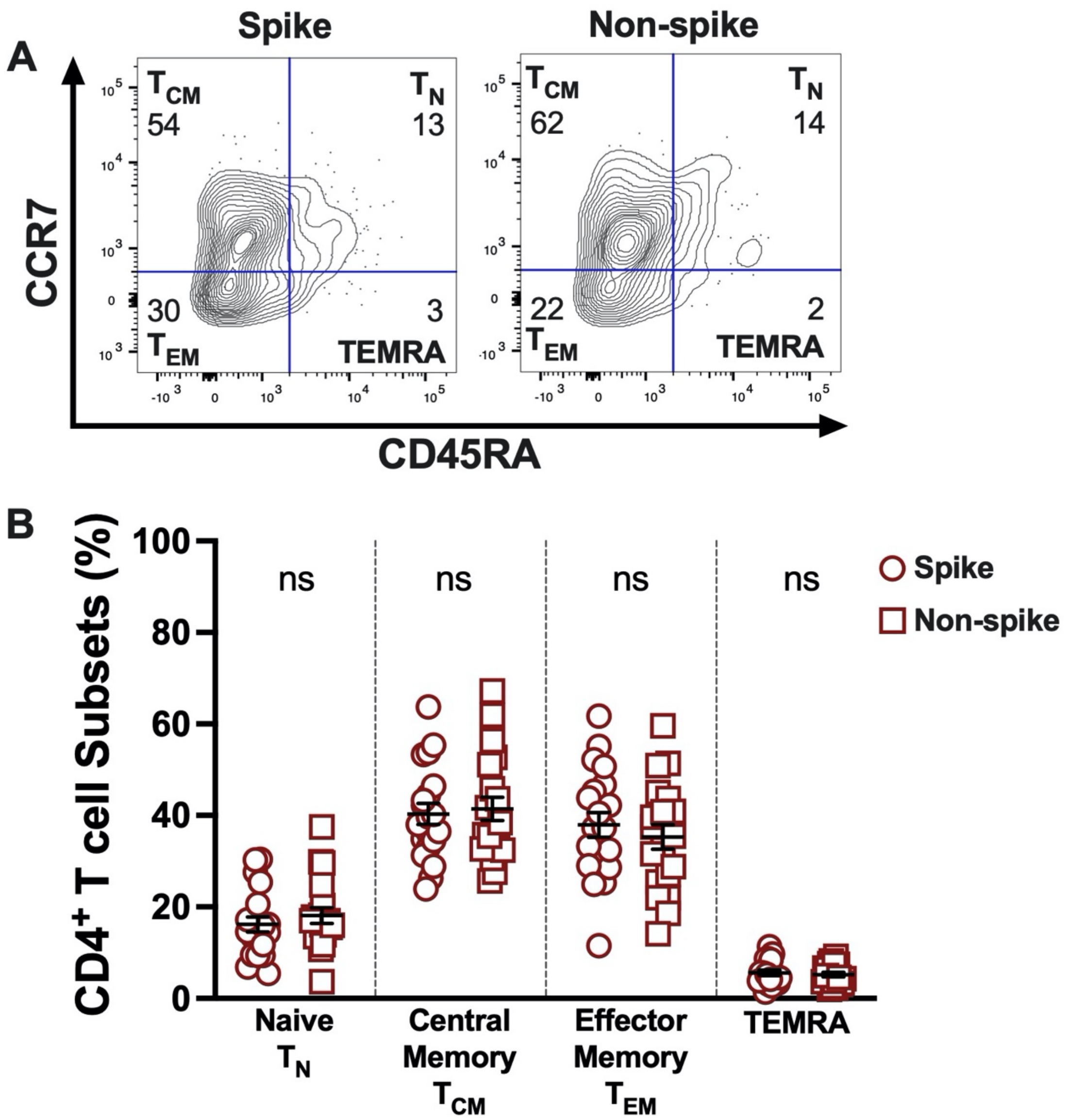


**Supplementary Figure 3. Memory phenotype of SARS-CoV-2-specific CD4$^{+}$ T cells in recovered COVID-19 patients**. The memory phenotype of the antigen-specific CD4$^{+}$ T cells was determined in the COVID-19 recovered subjects (n=20). The PBMCs were stimulated with the peptide megapool specific to Spike glycoprotein (Spike) or to the remainder of the SARS-CoV-2 polyprotein (Non-spike). (**A**) Example FACS plot for analysing the memory phenotype on antigen-specific AIM$^{+}$ (OX40$^{+}$CD137$^{+}$) CD4$^{+}$ T cells. (**B**) Proportion of memory phenotype in the antigen-specific AIM$^{+}$CD4$^{+}$ T cells. Phenotypes of AIM$^{+}$CD4$^{+}$ T cells were defined by the expression of CD45RA and CCR7. CD4$^{+}$ T cell subsets were defined as naïve ($T_N$) cells (CD45RA$^{+}$CCR7$^{+}$), central memory ($T_{CM)}$ cells (CD45RA$^{-}$CCR7$^{+}$), effector memory ($T_{EM}$) cells (CD45RA$^{-}$CCR7$^{-}$), and TEMRA cells (CD45RA$^{+}$CCR7$^{-}$). Black bars indicate the geometric mean. Statistical comparisons were performed by two-tail Mann-Whitney test. ns: non-significant.

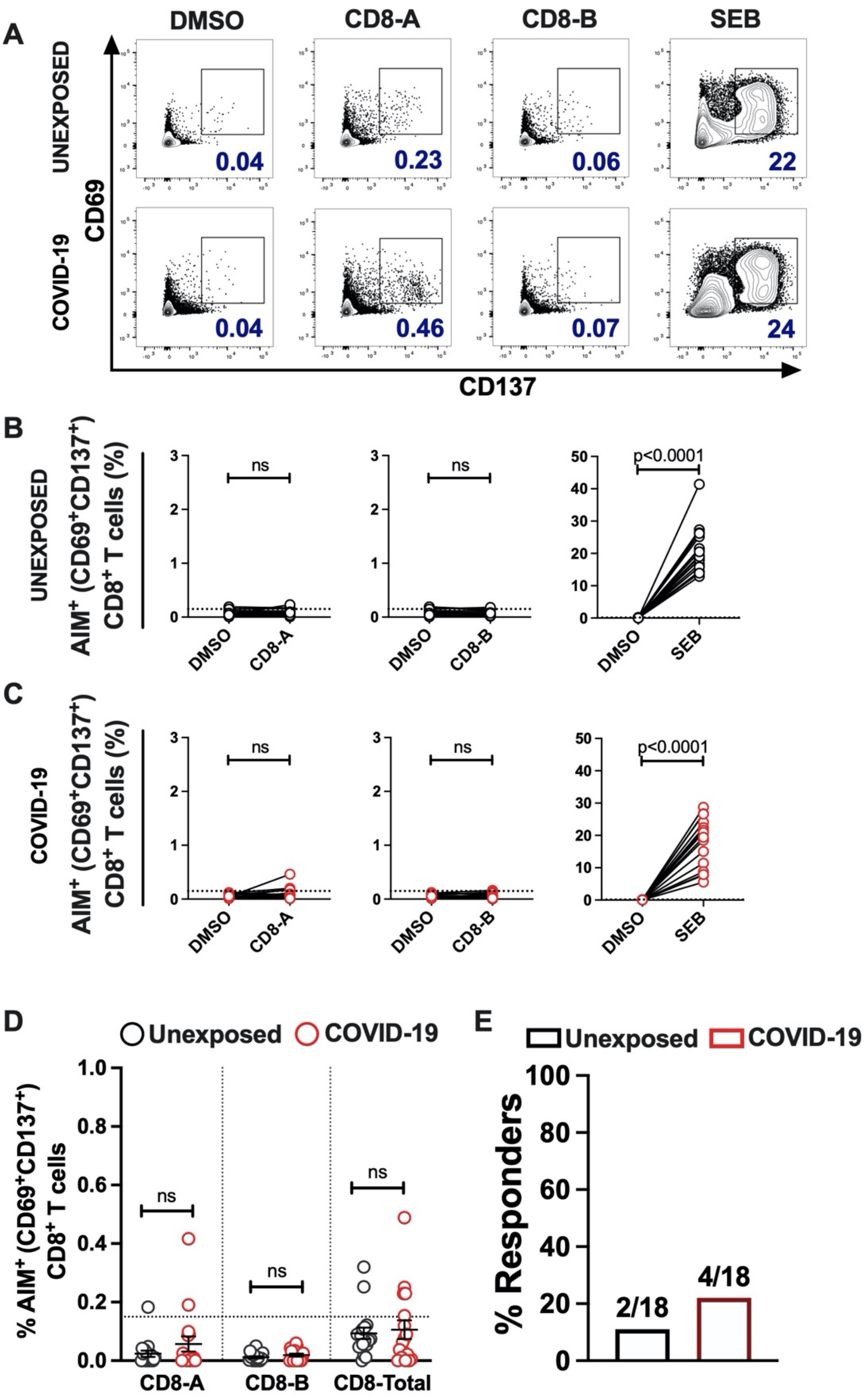


**Supplementary Figure 4. SARS-CoV-2-specific CD8$^{+}$ T cells response in unexposed donors and recovered COVID-19 patients.** The magnitude of SARS-CoV-2 specific CD8$^{+}$ T cells was determined in PBMCs collected from unexposed donors ("Unexposed", n=18) prior to pandemic and in COVID-19 patients ("COVID-19", n=18) up to 5 months of recovery. The PBMCs were stimulated with the class I peptide megapool (MP) comprises of 628 predicted CD8 epitopes and split into two MPs (CD8-A and CD8-B). DMSO was used as the negative control, and SEB was used for positive stimulation control. **(A)** Representative FACS contour plots of unexposed and COVID-19 patient in stimulation conditions of DMSO, CD8-A MP, CD8-B MP, and SEB. Paired graphs depicting the reactivity of AIM$^{+}$ (CD69$^{+}$CD137$^{+}$) CD8$^{+}$

T cells between the negative control (DMSO) and antigen-specific stimulation in (**B**) Unexposed donors (**C**) COVID-19 patients. **(D)** Percentage of the SARS-CoV-2 specific $AIM^+$ ($CD69^+CD137^+$) $CD8^+$ T cells after stimulation with the class I MPs (CD8-A, CD8-B) and the combined data of both the MPs, analysed in the same samples as in panel B and C. The data obtained in MPs stimulation were background subtracted with the respective DMSO negative controls. Black bars indicate the geometric mean. **(E)** Frequency of responders in unexposed and COVID-19 recovered subjects determined from the combined data of CD8-A and CD8-B MPs after background subtracted from the DMSO negative controls. The value on bars denote the number of responders/total number of donors tested. Dotted line in panels B, C and D represent the limit of detection. Statistical comparisons were performed by (B-C) Wilcoxon paired t-test and (D) two-tail Mann-Whitney test. ns: non-significant.
